# Air cleaning reduces incident infections in day care - an interventional crossover study

**DOI:** 10.1101/2024.09.25.24314350

**Authors:** Ville Vartiainen, Inga Ehder-Gahm, Johanna Hela, Anni Luoto, Jussi-Pekka Juvela, Petra Nikuri, Aimo Taipale, Natalia Lastovets, Sampo Saari, Ilpo Kulmala, Arto Säämänen, Enni Sanmark, Piia Sormunen

## Abstract

**Background:** While possibility of airborne transmission in the spread of common respiratory infections, there is no consensus on the relative importance of airborne infection route in real-life. This study aimed to investigate the significance of the airborne transmissions and the effectiveness of air cleaning in reducing infections among children in daycare.

**Methods:** A cross-over study was conducted in four daycare centers in Helsinki. All children attending the daycare were invited to participate (n = 262) and the sole inclusion criterion was that the children were expected to stay in the same day care center for the two-year duration of the study. 51 subjects were included in the final analysis. Clean air flow rate was increased by 2.1-2.9 times compared to baseline mechanical ventilation of the premises. The effect of intervention was assessed using negative binomial regression.

**Results:** The intervention reduced incident infections from 0.95 to 0.78 infections per child per month among the children (primary outcome) in daycare. The reduction attributed to intervention in the statistical model was 18.0 % (95% CI 2.1-31.3 %, p = 0.028).

**Conclusions:** We observed a significant decrease in incident infections without implementing any other infection mitigation strategies but air cleaning. Our results challenge the current paradigm which emphasizes fomite and contact transmission and infection control measures that target these pathways. As ventilation and air cleaning can only affect particles able to float in the air stream, our results support the significance of airborne transmission among common respiratory pathogens as well as air cleaning as an infection control measure.

## Introduction

Prior to the Covid-19 pandemic, it was generally accepted within the scientific community that the predominant modes of transmission for common respiratory viruses were through droplets or direct contact, while aerosol transmission was considered primarily associated with specific pathogens such as tuberculosis or measles, or specific high risk conditions.^1^ At that time, droplets were simply defined as particles exceeding five micrometers in diameter and tought to settle within a radius of less than one meter.^2^ Conversely, airborne transmission was defined as the dissemination of droplet nuclei that remain infectious and suspended in the air for extended periods and over greater distances.^2^ However, the surge in research during the Covid-19 pandemic significantly revised the prevailing understanding of aerosol classification and the transmission mechanisms of respiratory infections.^3,4^ Correspondingly, the growing evidence highlights the possibility of airborne transmission in the spread of common respiratory infections.^5,6^ However, there is no consensus on the relative importance and role of airborne infection in real-life. While WHO no longer promotes strict division between aerosols and ballistic droplets or any specific size of infectious respiratory particles, it still does not recommend adequate measures to control or prevent the spread of airborne.^7,8^ Instead it has called for better evidence on transmission routes and mitigation strategies of common respiratory pathogens.^8^

In natural settings, the significance of the airborne transmission can be demonstrated by examining interventions that specifically limit only aerosol transmission such as ventilation and air cleaning. Increased mechanical ventilation leads to greater dilution and removal of aerosols, resulting in lower airborne pathogen concentrations. It is also shown that ventilation has only a minimal effect on large ballistic droplets that do not remain suspended in the air stream.^9^ There is also similar evidence for the effectiveness of air cleaners They have been demonstrated to significantly reduce particle numbers^10^ and air filters equipped with High Efficiency Particulate Air (HEPA) filtration technology have been shown to decrease the presence of SARS-CoV-2 and other bioaerosols in air samples^11^. Air cleaners are proposed as a quick solution for spaces where the ventilation is insufficient. Although the effectiveness of ventilation and air cleaners in reducing the risk of infectious diseases has been discussed^12–14^, high-quality controlled intervention studies demonstrating the benefits of air cleaners or effective ventilation are lacking.

Respiratory infections are the most prevalent among children in early childhood education, with numerous episodes often occurring within a single year. ^1^ However, common prevention and control measures such as masks, social distancing, and hand hygiene are implemented inadequately or not at all among children. Thus, previously prevention and control have primarily been based on absences during illness.^15^ For these reasons, daycare provides an excellent platform for investigating the relative importance of airborne transmission of common infection diseases in a real-life setting. Concurrently, illnesses among daycare children also have significant health and economic impacts on society, necessitating effective infection control measures. ^15^

In this study, we present the results of a multidisciplinary effort by medical professionals, aerosol physicists, and building system engineers aiming to assess the added benefit of air cleaning on common respiratory infections in buildings with modern mechanical ventilation and to explore its implications on the transmission routes of these diseases.

## Materials and Methods

The study was conducted in four day care centers in Helsinki, Finland. Day care centers served between 85 - 123 children each, all aged one to six years and the staff comprised 19 to 21 members. Due to nature of this study, randomization on an individual level was not possible but the day care centers were randomized in two sequences with day cares A and B in Intervention-Control sequence and day cares C and D in Control-Intervention sequence. The study included two periods 11/22-4/23 and 11/23-4/24. Children expected to attend the whole duration of the study were invited to participate and the recruitment was conducted during October 2022. There were no exclusion criteria. Subjects who withdrew their consent or did not answer questionnaires during both winters of the study were excluded from the final study population (FIG 1). The sample size was calculated as described by Lui for AB/BA crossover using Poisson regression with α = 0·05 and β = 0·80.^16^ The number of infections in control group was assumed to be 10 and RR 0·9. With estimated drop out rate of 20% the required sample size was estimated to be N = 188.

**Figure 1:**
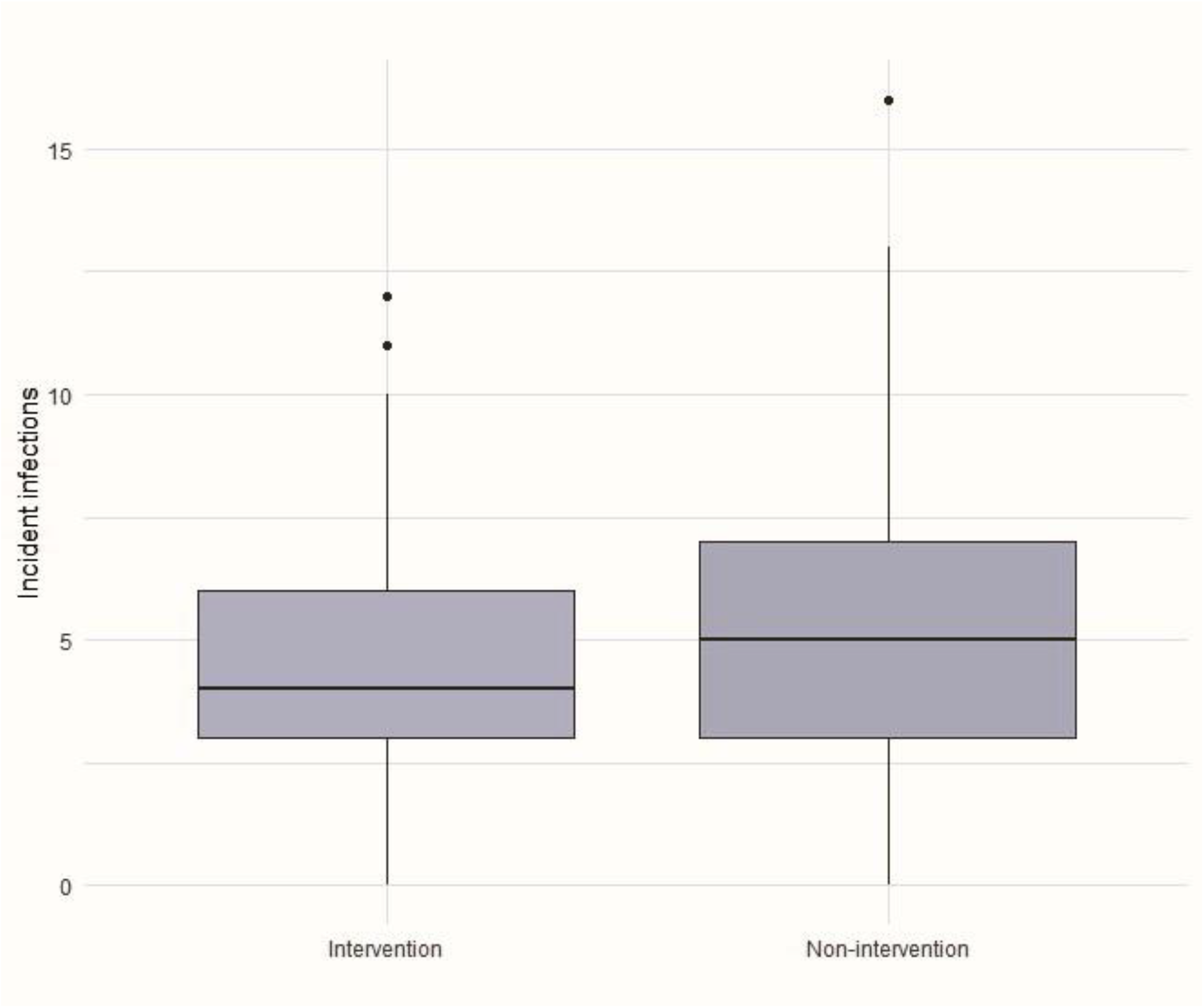
Cumulative incident infections during the intervention and non-intervention periods of the study.

The day care centers were built in 2001, 2002, 2009, and 2013 and were equipped with modern mechanical ventilation systems meeting with current building code. The ventilation systems of all buildings were examined and maintenance prior to the initiation of the study. The ventilation systems air handling units were equipped with heat recovery (no recirculation air) and were operated during working days from 4:00 am to 6:00 pm in all buildings. The day care centers were located in the same district in the city of Helsinki.

The intervention consisted of a total of 45 portable room air cleaners (PAC) alongside the existing ventilation systems. Most of the air cleaners were tested in the laboratory applying ANSI/AHAM AC-1-2020 test standard to determine their particulate clean air delivery rates (CADR). The CADR values are used as a proxy for clean air flow rate for PAC. The supply air flow from ventilation, the occupancy rate and the usage of the room were investigated through a survey when sizing air cleaning. The placement of PAC aimed to achieve the maximum benefit in reducing infections.^14^ The used air cleaners are described in detail elsewhere.^15^ The used CADR values per PAC varied between 125 and 1500 m3/h. All air cleaners were operated from 6:00 am to 4:00 pm, coinciding with the activities in the day care centers. The operation of the devices was monitored with smart plugs. The air flow rates of each day care center are presented in table 1.

**Table 1.** Clean air flow rates in day care centers.

| Day care center | Number of persons | Clean air flow rate (l/s/person) |  | Multiplication factor in clean air flow rate |
| --- | --- | --- | --- | --- |
|  |  | Control sequence (Mechanical ventilation only) | Intervention sequence (Mechanical ventilation + PAC) |  |
| A | 123 | 12.8 | 27.5 | 2.2 |
| B | 110 | 11.9 | 29.1 | 2.4 |
| C | 105 | 19.0 | 40.6 | 2.1 |
| D | 105 | 14.5 | 42.3 | 2.9 |
| **Average** |  | **14.5** | **34.9** | **2.4** |

Clean air flow rates of all day care centers during the control sequence (existing mechanical ventilation only) and intervention sequence (portable air cleaners (PAC) alongside the existing mechanical ventilation). Day care center averages are based on occupancy weighted means of the spaces in the day care.

Children’s illnesses were the primary outcome and parents’ absence from work the secondary outcome of the study. The information was collected through weekly electronic surveys answered by parents. The analyzed questions in the diary were “Has your child been ill during the previous week? (yes/no)” and “Did the child’s illness cause work absences to the adults?” Negative binomial regression models were constructed to study the effect of the intervention. Incidence infections and adult’s absences were included as counts, age as continuous, and sex and period as categorial variables. Subject ID was included for random effects. Statistical analyses were done using R version 4.3.3.

The study was registered at clinicaltrials.gov (NCT05569330). It was approved by the ethical committee of Helsinki and Uusimaa (HUS/14231/2022) and was conducted according to the Declaration of Helsinki.

### The role of funding

The funders of this study did not have any role in planning the study, interpretation of the results or writing of this manuscript.

## Results

25 females and 26 males of age between 1-5.9 (mean 3.9) years were included in the final study population (fig. 1). Mean response rate to the surveys was 63.2% (range 45.8-80.6%). In addition to intervention, age and sex of the subject and intervention period were also included in the analysis. The effect of age or sex were not statistically significant, nor did they affect the other estimates and were, therefore, left out of the models. The effect of period was statistically significant but its absence or presence in the model did not alter the estimate for the intervention.

In the statistical model the estimate for the effect of the intervention on incident infections among the children in daycare (primary outcome) was 0.199 (95% CI 0.02-0.37, p = 0.02) resulting in 18.0 % (95% CI 2.1-31.3 %, p = 0.028) reduction of infections. The estimate for the period was -0.229 (95% CI -0.406–0.052, p = 0.011). The number of incident infections during intervention and non-intervention periods is presented in figure 2. The total number of recorded absences was 239 during intervention periods and 292 during non-intervention periods resulting in 0.78 and 0.95 incident infections per child per month during intervention and non-intervention periods, respectively. The secondary outcome was work absences of the parents due to these infections. While we observed a decrease of 15·8 % it did not reach statistical significance (95% CI -0.29 – 0.64, p = 0.46).

## Discussion

Daycares are notorious for compromised hand hygiene, high exposure to ballistic droplets due to poor coughing etiquette and maximized fomite transmission routes for infections. Consequently, protection against infections has traditionally relied on absence during illness and building immunity through exposure.^16^ In this study, we demonstrated a clinically and statistically significant reduction in incident infections in daycares due to significant increase in clean air flow rate without limiting any other transmission route. The intervention was effective even though the subjects spend only part of their time in the day care and there are numerous other sites for infections to transmit. Our results strongly suggest that contrary to the previous paradigm of the role airborne transmission it plays an important role in transmission of common infections.^8^ As a result, the importance of ventilation and air cleaning in the prevention and control of common infections should be emphasized in guidelines and recommendations. The secondary outcome of adults’ absences did not reach statistical significance. This may be confounded by the fact that many children in day care commonly have younger siblings in home care and thus, the illness does not cause parent to be absent from work. Our results are in line with previous literature as the trend between the child’s illness and parent’s absences generally follow the same trend, but there were fewer parental absences from work than sick days taken by children.^17,18^

The air cleaning has been earlier observed to decrease the number of colony-forming units in intensive care units. Additionally, a correlation has been reported between pathogens detected in the air of intensive care units and hospital-acquired infections.^19^ In contrast, Falkenberg et al did not observe the effect of HEPA filters on COVID-19 incidence during the Omicron wave in day care units in Germany.^20^ The study, however, has several methodological limitations: part of the data was collected retrospectively, the study lasted only one year and no cross-over methodology was used, children’s demographic data were not considered, and any information on dimensioning, positioning or even running the air cleaning equipment were not reported.

Particles are generated in all respiratory activities including tidal breathing and speaking.^21^ Since respiratory infectious particles originate from the fluid lining the mucosa, any viruses replicating at the site of aerosol generation can be contained within the emitted aerosols. For example, in the case of SARS-CoV-2, even tidal breathing has been shown to generate aerosol particles that carry viable viruses and copies of SARS-CoV-2 RNA.^22^ Therefore, it is not surprising that also the common respiratory pathogens transmit through the air. However, the association between airborne transmission and illness has not been previously demonstrated, and for example, in recent challenge studies, study subjects have been infected with nasal drops.^23,24^ Since air cleaning has shown only a minimal effect on the presence of ballistic droplets and fomite route cannot be controlled in the interaction of children under 6 years old in daycare our study demonstrates a strong association between air cleaning and reduced infections, highlighting the significant role of airborne transmission in common infections.^9,16^

Air flow rates of mechanical ventilation are typically designed for good indoor air quality under normal conditions and fresh, outdoor air flow rates of 8–10 l/s/person are common. However, with the increase in the number of infectious particles these rates may be inadequate for controlling airborne infection transmission. Recommendations for improved ventilation during COVID-19 suggest significantly higher air flow rates, ranging from 14 l/s/person to 20-25 l/s/person.^25,26^ In this study, the control case outdoor air flow rates varied between 11.9 l/s/person to 19.0 l/s/person. During the intervention, additional portable air cleaners increased the clean air flow rates up to 27.5 - 42.3 l/s/person multiplying them 2.1-2.9 times. It is, however, important to note that this study was conducted in buildings with modern mechanical ventilation meeting the current building code requirements. Still a significant decrease in incident infections was observed due to the air cleaning. This emphasizes the crucial role of indoor air purity in infection prevention also in modern buildings in the future.

It is important to acknowledge several limitations in this work. In this study the intervention was aimed to the environment rather than directly to the participants. Therefore, randomization within the day care centers was not possible and had to be done on day care center level rather than individual level. We were unable to find a solution to adequately implement placebo control in this study. Using only fans to move the air still causes significant mixing and potential dilution of the pathogen concentration in air possibly affecting the infection risk.^13^ As all cross-over trials this one is also subject to carry over effects. Although unlikely, it is possible that the participants in intervention-control sequence did not develop similar immunity to common respiratory pathogens as participants in the control-intervention sequence due to reduced infection rate and thus had more infections during the control period. The two periods included in the study were different in terms of infections most likely due to natural variation of seasonal epidemics which is demonstrated e.g., in national sewage surveillances.^28^ However, as controlling for period effect in the statistical model did not alter the effect of intervention, we concluded the possible bias was adequately controlled. While cross over design is well controlled for confounding factors, we did fall short from our recruitment goal which prevented us from conducting subgroup analysis. The effect size was higher than originally anticipated and therefore, statistical power was sufficient despite the problems in recruitment. We also had a significant number of dropouts during the study, but they were distributed evenly between the groups and are not likely to bias the results. As the day care units in this study were deemed to be typical and the buildings had modern mechanical ventilation, we expect the results to generalize well. However, in buildings with outdated ventilation systems the relative effect of PACs is likely to be bigger.

## Conclusions

In this work we have presented the effect of air cleaning in reduction of common infections in day care units. We observed a significant decrease in incident infections without intervention to any other transmission routes or implementing other infection mitigation strategies. Our results challenge the current paradigm which does not acknowledge the important role of airborne transmission of common infections and emphasizes fomite and contact transmission particularly in the case of prevention and control measures. As ventilation and air cleaning can only affect airborne particles, our study demonstrates the significance of airborne transmission of common respiratory pathogens and the crucial role of air cleaning in the prevention and control of these infections.

## Data Availability

The data will not be publicly shared due to Finnish legislation on medical research and statement of the ethics committee.

## Acknowledgements

Tero Vahlberg is acknowledged for expert statistical support. Riku Kivisaari is acknowledged for the initial idea and stimulus for this work. We would like to thank all the participating families and the staff of the day care centers for their participation and support for this study.

## Contributors

Ville Vartiainen conceptualisation, data curation, formal analysis, funding acquisition, investigation, supervision, validation, visualisation, writing – original draft, and writing– review & editing, methodology, directly accessed the data

Inga Ehder-Gahm conceptualisation, methodology, investigation, writing – original draft

Johanna Hela investigation, data curation, directly accessed the data, writing– review & editing

Anni Luoto methodology, investigation, writing– review & editing

Jussi-Pekka Juvela methodology, investigation, writing– review & editing

Petra Nikuri investigation, writing– review & editing

Aimo Taipale methodology, conceptualisation, investigation, writing– review & editing

Natalia Lastovets methodology, investigation, writing– review & editing Sampo Saari methodology, investigation, writing– review & editing

Ilpo Kulmala methodology, investigation, writing– review & editing

Arto Säämänen methodology, investigation, writing– review & editing

Enni Sanmark conceptualisation, funding acquisition, supervision, writing – original draft, and writing– review & editing, methodology, project administration, resources

Piia Sormunen conceptualisation, funding acquisition, supervision, writing – original draft, and writing– review & editing, methodology, project administration, resources

## Data sharing

The data will not be publicly shared due to Finnish legislation on medical research and statement of the ethics committee. Data, protocol, consent form, and all questionnaires are available from corresponding author upon reasonable request.

